# Quantitative assessment of head movement dynamics in dystonia using visual perceptive deep learning: a multi-centre retrospective longitudinal cohort study

**DOI:** 10.1101/2023.09.11.23295260

**Authors:** Robert Peach, Maximilian Friedrich, Lara Fronemann, Muthuraman Muthuraman, Sebastian R. Schreglmann, Daniel Zeller, Christoph Schrader, Joachim Krauss, Alfons Schnitzler, Matthias Wittstock, Ann-Kristin Helmers, Steffen Paschen, Andrea Kühn, Inger Marie Skogseid, Wilhelm Eisner, Joerg Mueller, Cordula Matthies, Martin Reich, Jens Volkmann, Chi Wang Ip

## Abstract

**Background:** Dystonia is a neurological movement disorder characterised by abnormal involuntary movements and postures, particularly affecting the head and neck. However, current clinical assessment methods for dystonia rely on simplified rating scales which lack the ability to capture the intricate spatiotemporal features of dystonic phenomena, hindering clinical management and limiting understanding of the underlying neurobiology. To address this, we developed a visual perceptive deep learning framework that utilizes standard clinical videos to comprehensively evaluate and quantify disease states and the impact of therapeutic interventions, specifically deep brain stimulation. This framework overcomes the limitations of traditional rating scales and offers an efficient and accurate method that is rater-independent for evaluating and monitoring dystonia patients.

**Methods:** To evaluate the framework, we leveraged semi-standardized clinical video data collected in three retrospective, longitudinal cohort studies across seven academic centres in Germany. We extracted static head angle excursions for clinical validation and derived kinematic variables reflecting naturalistic head dynamics to predict dystonia severity, subtype, and neuromodulation effects. The framework was validated in a fully independent cohort of generalised dystonia patients.

**Findings:** Computer vision-derived measurements of head angle excursions showed a strong correlation with clinically assigned scores, outperforming previous approaches employing specialised camera equipment. Across comparisons, we discovered a consistent set of kinematic features derived from full video assessments, which encoded information relevant to disease severity, subtype, and effects of neural circuit intervention more strongly and independently of static head angle deviations predominantly used for scoring.

**Interpretation:** The proposed visual perceptive machine learning framework reveals kinematic pathosignatures of dystonia which may be utilized to augment clinical management, facilitate scientific translation and inform personalised and precision approaches in Neurology.

**Research in context:** *Evidence before this study:* Clinical assessment of dystonia, a neurological movement disorder, has traditionally relied on rating scales that aim to simplify complex phenomenology into lower-dimensional rating items. However, these score-based assessments have significant clinimetric limitations and do not fully capture the rich spatiotemporal dynamics of dystonic phenomena, which are crucial for clinical judgment and pathophysiological understanding. In contrast, recent investigations in animal models of dystonia have already demonstrated the utility and relevance of quantitative methods for phenotyping, which gradually supersedes previous observer-dependent behavioural analyses. Taken together, this has led to a need for more objective and detailed clinical evaluation methods of dystonia. We performed a PubMed search up to July 2023 combining the terms “dystonia” AND (”deep learning” OR “machine learning” or “computer vision” OR “vision-based” OR “video-based”) AND (”angle” OR “kinematic” OR “rating” OR “scoring” OR “movement analysis”) including abstracts in English or German. The search yielded three studies that validated vision-based frameworks for automating the assessment of cervical dystonia severity compared to clinician-annotated ratings. Two of these studies focused on deriving head angle deviations from specialised camera setups, while the third study utilised computer vision in a retrospective video dataset recorded using conventional equipment. These studies reported fair to moderately strong correlations between vision-based head angle measurements and clinical scores. Additionally, two studies investigated computer vision for assessing head tremor in the context of cervical dystonia: one single case report demonstrated the clinical validity of computer vision-derived head angle and head tremor metrics, while a retrospective cross-sectional study reported moderately strong clinical agreement of computer vision-derived head oscillation metrics across different dystonia subgroups. Two additional studies used computer vision-based kinematics to quantify dystonia-like phenomena in rodent models of monogenetic dystonia, demonstrating utility in both phenotype and genotype predictions. However, most of the clinical studies were limited to static task conditions, where patients attempted to hold a neutral position of the head, thus not providing a naturalistic account of dystonia. Moreover, beyond head angular deviations and oscillation metrics, no study explored a broader kinematic feature space that reflects the true spatiotemporal complexity of dystonic movements. Additionally, the studies assessed patients at single time points without considering different therapy conditions, particularly the effects of deep brain stimulation, which is a highly effective intervention targeting brain circuits. Nor did they compare dystonia sub-types, such as cervical and generalised systonia.

*Added value of this study:* In this study, we present a comprehensive visual perceptive deep learning framework that addresses the gaps in current dystonia assessments. We use this framework to retrospectively analyse a unique dataset from three multi-centric, studies encompassing video examinations of patients along the dystonic severity continuum, including different deep brain stimulation states. Our framework goes beyond the automation of suboptimal symptom severity assessments by reverse engineering a set of clinically inspired kinematic features. The resulting high dimensional, yet intuitively interpretable kinematic feature space enabled us to explore disease states and effects of brain circuit therapies in a level of detail comparable to experimental neuroscientific investigations. Through a data-driven approach, we have identified a consistent set of only four dynamic parameters that encode dystonia severity, subtype, and the efficacy of brain circuit interventions. Notably, these features are independent of static head angle deviations, which play a central role in dystonia severity scores, pointing to the involvement of partially distinct neurobiological processes not captured by these scores. Our findings align with emerging concepts of symptom-specific brain circuits and findings in rodent models of dystonia, thereby exemplifying the visual perceptive framework’s potential to augment clinical management and bridge translational gaps in movement disorders research. By providing a more comprehensive and precise assessment of the disorder, our study offers valuable insights for improved treatment strategies and further understanding of dystonia’s complex neurobiology.

*Implications of all the available evidence:* The available evidence collectively underscores the limitations of traditional rating scales in capturing the informative spatiotemporal dynamics of dystonic movements, emphasizing the need for more objective and granular evaluation methods. In line with recent animal studies using computer vision for dystonia quantification, recent clinical studies have shown the potential of computer vision-based frameworks in automating cervical dystonia severity assessment and capturing head tremor metrics. However, their underlying study designs may inadvertently reinforce limitations associated with the clinical scoring process. In this study, we introduce a comprehensive visual perceptive deep learning framework that serves as a powerful platform to augment clinical judgement and generate valuable pathophysiological insights by extracting a set of clinically inspired, interpretable kinematic features. Our findings have implications beyond dystonia, showcasing the utility of visual perceptive frameworks in enhancing clinical management and fostering integration with advanced neuroimaging and neurotechnological methods. This study opens doors for future translational research to explore the broader application of computer vision and deep learning techniques to derive kinematic signatures of movement disorders across species and experimental conditions, promising more precise and personalised assessments that can significantly improve therapeutic strategies and patient outcomes.

## Introduction

Dystonia is a neurological disorder characterised by abnormal movements and postures caused by involuntary muscle contractions[1]. It is recognised as the third most prevalent movement disorder, with recent estimates as high as 732 per 100,000 individuals [2]. Despite advancements in understanding the epidemiological, neurogenetic, and neurobiological factors associated with dystonia, the identification of objective biomarkers remains challenging. Consequently, the diagnosis, monitoring of treatment outcomes, and classification of dystonia heavily rely on clinical phenomenology. This entails considering various factors, such as the distribution of affected body regions, which allows for categorising dystonia along a severity spectrum of focal, segmental and generalised manifestations[2]. However, dystonic movements exhibit highly complex spatiotemporal characteristics, involving a combination of tonic and phasic elements, such as twisting, tremulous oscillations, and overflow to other body regions, occurring on variable time scales and exacerbated or alleviated by certain movements [1, 3–5]. Achieving precise clinical phenotyping of dystonia poses a significant challenge, demanding expert visual perception skills[6].

To accurately assess disease progression and therapeutic outcomes in dystonia, it is essential to employ reliable and well-defined operational measures that can be consistently measured and interpreted across diverse clinical settings and practitioners. This is of particular relevance for assessing outcomes of available therapies, ranging from oral medication to Botulinum neurotoxin injections for selective muscle weakening and deep brain stimulation (DBS)[7]. To date, clinical rating scales such as the Toronto Western Spasmodic Torticollis Rating Scale (TWSTRS) for cervical dystonia and the Burke-Fahn-Marsden Dystonia Rating Scale (BFMDRS) for generalized dystonia have been extensively utilised for this purpose [8–10]. These scales aim to condense complex clinical observations into simplified representations, relying on a limited set of categorical items, such as head-angle deviations in attempted neutral head position, encoded by a few ordinal values. Although this simplification offers advantages in time-sensitive clinical settings, it is accompanied by significant clinimetric limitations, including substantial inter-rater variability[11–13]. Furthermore, the original versions of these scales fail to quantify important information regarding abnormal movement trajectories, action-induced changes of dystonia, dystonic overflow (i.e., the spread of dystonic posturing/movement to adjacent body parts), and tremor, which has recently been recognised as affecting a majority of dystonia patients [14]. Yet, emerging evidence from animal models highlights the critical role played by the rich spatiotemporal structure of motor behavior in understanding the pathocircuitry of dystonia, thereby shaping our approach to investigation and treatment [15–17]. The lack of standardised operational and shared measures hampers translational efforts, thus necessitating the development of objective outcome measures[3, 18].

To address the challenges of dystonia assessment, researchers have explored various instrumented solutions, such as electromyography[7] or body-worn sensors[19]. However, the successful integration of these approaches into clinical practice has proven elusive[20]. Contactless, vision-based methods utilising multiple and/or special depth cameras have shown promise in extracting head angles in cervical dystonia. Nevertheless, their clinical validity has been limited, especially when operating under monocular conditions [21, 22]. In this context, computer vision, a branch of contemporary artificial intelligence, has emerged as a disruptive and promising technology in clinical neuroscience and broader medical applications [23–26]. By leveraging convolutional neural networks (CNNs), visual perceptive frameworks offer several advantages, including real-time 3D human pose tracking derived from monocular 2D videos captured by consumer-grade camera hardware [27, 28]. These advancements have significantly improved head pose estimation [29–31], some of which have been employed to semi-automate TWSTRS ratings [20]. However, these studies have primarily focused on reproducing the rating score by quantifying static head angular deviations in a single fixed head position, thereby reinforcing the aforementioned limitations and biases associated with the rating scale. Our hypothesis is that a naturalistic approach, which incorporates both gestalt aspects and the dynamics of head movement, will lead to a more accurate and ecologically valid assessment of dystonia. This approach will enable us to capture subtle variations and intricate patterns that may have been overlooked by previous constrained methods. Furthermore, we propose that including healthy controls and different dystonia subgroups, with repeated recordings at various therapeutic states (e.g., different DBS settings), will allow us to explore multiple facets of specificity in these digital physiomarkers.

In this study, we have developed a visual perceptive deep learning framework that utilises computer vision to analyze the dynamics of natural head movement. The goal was to identify distinct patterns, or pathosignatures, that have diagnostic and therapeutic implications. By doing so, we aimed to enhance our understanding of the underlying pathophysiology of dystonia and effects of therapeutic neuromodulation. Specifically, we trained a novel convolutional neural network to predict movement states, and we combined the outputs with head angles obtained from a benchmark algorithm, MediaPipe, to extract both static and kinematic features from patients undergoing clinical dystonia examinations. To demonstrate the feasibility of our approach, we conducted a retrospective cohort study to assess the agreement between predicted severity and clinical ratings, establishing how both static and dynamic variables change in response to DBS. Subsequently, we validated the predictive accuracy of dynamic variables using an additional cohort of patients with generalised dystonia. Lastly, we provide insights into the added value of the dynamic variables in differentiating between patients with cervical dystonia and those with generalised dystonia.

## Methods

### Study design and participants

We sourced clinical video data documenting the severity of cervical and generalised/segmental dystonia from two prospective, longitudinal, multi-centre cohort studies investigating the therapeutical effect of pallidal DBS on dystonia [32–34] and a third, multi-centre retrospective investigation analyzing clinical outcomes using advanced neuroimaging techniques [35]. The sourced data was split into two data sets, based on dystonia subtype: (i) cervical and (ii) generalised dystonia. The cervical dystonia cohort comprised 86 cervical dystonia patients from Rostock, Heidelberg, Dusseldorf, Berlin, Innsbruck, Oslo, Hannover, Kiel, Würzburg. The generalised dystonia cohort, used for independent validation, comprised 30 patients from the same centres. Individual datasets were included if (i) they contained at least one pre-operative clinical rating video showing the full dystonic phenotype and a video from the chronic postoperative phase (3-15 months post surgery) documenting the effects of clinically programmed DBS and (ii) both videos fulfilled minimal criteria ensuring video quality, which were chosen to reflect the current best practice in clinical computer vision approaches [20, 23, 24]. These were: (i) front view perspective of a single individual sitting on a chair, (ii) no significantly obscuring items on patients (e. g. excessive head dressings with externalised DBS device), (ii) no excessive camera movements, variable zoom depths or lighting insufficient to identify typical body landmarks (e.g., eyes), (iii) continuous presence of head and neck in the camera frame. A final set of 232 videos, comprising a total of 116 individual patients, was analysed in this study. All videos were recorded with standard consumer grade camera hardware, in most instances mounted on a tripod. The minimal spatiotemporal resolution was 540 *×* 540 pixels and 24 frames per second. An additional cohort of 22 healthy controls underwent a structured TWSTRS examination and a head position matching task. This task was precisely timed to map ground truths of head movement range along each of the three principal rotational axes (pitch, yaw, tilt; see supplementary Figure S1 for the detailed protocol).

#### Ethics approval

This study was approved by the Julius-Maximilians University ethics committee (AZ 301/20). The original studies had been approved by the responsible ethics committees.

#### Clinical scoring

Respective dystonia severity rating scales, i.e., Burke-Fahn-Marsden dystonia rating scale (BFMDRS) for generalised dystonia and the Toronto Western Spasmodic Torticollis severity part (TWSTRS) for cervical dystonia, had originally been administered in an open-label approach or by one expert rater. In order to eliminate potential scoring biases and to extend the clinical rating to include head tremor[9], all videos were re-scored. To this end, video segments in which patients were asked to let their head drift to its natural null position were annotated. These segments partly reflect the individual dystonic phenotype and its severity (corresponding to TWSTRS severity element I). Three raters, two blinded senior movement disorders experts (DZ, CWI) and one junior investigator (LF) specifically trained using the TWSTRS teaching tape[8], applied the TWSTRS severity part. Three raters, two blinded senior movement disorders experts (DZ, SRS) and one junior investigator (LF), also applied an additional head tremor subscore from TWSTRS-2[9]. For subsequent analyses, we mainly focused on TWSTRS severity item assessing the time-weighted deviation of head posture from neutral straight ahead along three main rotational axes, namely pitch for antero-/retrocollis, yaw for torticollis and tilt for laterocollis. The original TWSTRS contains further items, which however failed to meet criteria for utility in subsequent investigations[9]. Each item is scored on an ordinal scale from 0 *−* 3 (laterocollis, anterocollis, retrocollis) or 0 *−* 4 (torticollis, head tremor), corresponding to increasing angle deviations of the head from the midline or in case of tremor, its amplitude, duration and dominant direction. Assessors’ ratings were collapsed into one ‘mean score’ for subsequent model evaluations.

### Visual perceptive deep learning framework

We built a comprehensive framework for assessing dystonia phenotype and severity, enabling automated kinematic evaluation directly from video. Our approach involved combining the outputs of two convolutional neural networks: one tracking facial landmarks and the other one for extracting gestalt information, represented as movement states. From each video, utilizing the deep learning outputs, we derived static variables during periods when patients were instructed to allow their heads to drift to a neutral position (referred to as the null position). In addition, dynamic variables capturing the patients’ natural movement patterns were extracted using the entire duration of the TWSTRS video examination. We should note that these clinical examinations often didn’t follow the full TWSTRS protocol, nor did they necessarily follow the prescribed ordering of movements.

To achieve face and head tracking, we utilized a pre-trained model from MediaPipe [30]. We opted for MediaPipe due to its real-time applicability and compatibility with mobile devices, which holds potential for point-of-care applications. The default tracking values of MediaPipe’s video mode (detection confidence: 0.5; tracking confidence: 0.5) were employed. Head angles were calculated relative to a neutral face-forward position along three axes of movement (torticollis, laterocollis, and antero-/retrocollis) using the face mask. We employed the orthogonal Procrustes technique to compute the rotation necessary to minimize the discrepancy between the rotated 3D face mask and a face-forward face mask, thus obtaining accurate head angles [36].

To track gestalt patterns throughout the videotaped examinations, which lacked a fixed protocol and order, we aimed to classify the head movement states on a frame-by-frame basis along the three principal axes. For this purpose, we developed a custom model trained on videos of healthy controls. We fine-tuned a pre-trained resnet50 convolutional neural network model in PyTorch for 30 epochs to achieve loss convergence. The training and validation data sets consisted of images from 22 healthy controls and 23 cervical dystonia patients, with participants exclusively assigned to one data split. 15 healthy controls and 15 patients were used for training, and the remainder for validation. Movement states (e.g., ‘face forward’ or ‘tilt left’) were labeled by a junior movement disorders expert (MF). The custom model demonstrated training and validation accuracies of 83.8% and 84.6%, respectively. We employed multilabel classification with a binary cross-entropy loss function during model training, and additional details are provided in the appendix.

Using the outputs of the two convolutional neural network models, we engineered several kinematic features that capture the temporal evolution of patients’ head trajectories beyond simple angular deviations. These kinematic features aimed to quantify clinically relevant observations in dystonia that are commonly noted but seldom quantified in clinical settings, such as movement overflow to other bodyparts as well as action-induced changes of dystonia, both resulting in asymmetrical or abnormal movement trajectories, dystonic tremor[14] and the complexity of dystonia characterised by the involvement of multiple axes in phasic or tonic movements and movement predictability over time. The features were partially harmonised with kinematic features recently reported to be relevant to dystonia phenotype and genetics in rodent models of dystonia [15, 16] as well as the characterisation of brain dynamics more broadly [37, 38]. The derived features primarily included correlations, symmetries, oscillatory and entropy-related characteristics, which are further described below.

#### Correlation features

The movement state predictions, represented by softmax outputs from the convolutional neural network, and the head-angle measurements (in degrees) are continuous values assigned to each frame of the video. To investigate the relationship between movement states, we calculated the Pearson correlation between them. By calculating the Pearson correlation between movement states, we are exploring the interdependence of different movement patterns. For instance, a high correlation between the prediction probabilities of rotation left and tilt left would indicate that the movement vectors blend or exhibit a certain degree of overlap when the patient rotates left. This suggests that the movement states become ‘entangled’ or ‘intermixed’ during specific actions, as recently suggested in experimental studies [15]. Healthy controls are expected to show minimal correlations between movement states and head angles, indicating precise and distinct control of head movements. These features aim to capture phenomena such as overflow and complexity, as well as abnormal movement trajectories.

#### Head oscillations

The primary frequency and amplitude of head-angle oscillations along each axis of motion were assessed using a Fourier spectrogram. To isolate the relevant oscillatory signals and remove intended head movements dictated by examination protocol, a bandpass filter with an order 6 Butterworth filter was applied, limiting the frequencies to the range of 2 *−* 10 Hz. These features aim to capture phasic characteristics, such as dystonic jerks and tremors. It is expected that healthy controls will exhibit minimal or no head oscillations in these frequency ranges.

#### Symmetry features

Each axis of head motion can exhibit movement in opposite directions from a neutral face-forward position. To quantify the symmetry of each motion axis, we calculated the proportion of time the head was oriented in one direction compared to the opposite direction. For instance, if a patient spent 7 seconds in rotation left and only 3 seconds in rotation right, the symmetry value would be calculated as (7 *−* 3)*/*10 = 4*/*10 = 0.4. Values closer to zero indicate a stronger symmetry, while large positive or negative values indicate a significant asymmetry. These features aim to capture fixed, tonically abnormal head deviations and asymmetrical movement trajectories. Healthy controls are expected to demonstrate a high degree of symmetry in their head movements.

#### Multi-scale entropy (MSE)

Entropy measures provide a quantitative way to assess the irregularity or complexity of time series data, making them well-suited for capturing the intricate and nonlinear dynamics often observed in dystonic movements [37, 38]. Abnormal movements in dystonia often exhibit both short-term irregularities (e.g., tremor) and long-term temporal patterns (e.g., sustained postures) that are not easily captured by traditional measures. MSE quantifies the complexity and regularity of dystonic movements at different temporal scales. By applying MSE to kinematic time series data, a scale-dependent measure of complexity can be obtained, potentially revealing specific temporal patterns or fluctuations associated with disease states or treatment effects. We hypothesis that DBS will increase the regularity and predictability of their movements, indicative of improved motor control.

### Evaluation of the visual perceptive framework

Performance in evaluating predominant direction and severity of dystonic head deviation was measured by Pearson correlation between the clinically annotated TWSTRS and the head angle excursion along each axis of motion respectively. Performance in evaluating the tremor component of patients was measured by Pearson correlation between the clinically annotated tremor score and the head angle oscillation amplitude along each axis of motion respectively. To measure the robustness of our approach, a validation dataset comprising generalised dystonia patients without clinical annotations was used. The same kinematic variables were extracted from the full videos and a statistical analysis comparing DBS conditions (preoperative off, postoperative on).

### Statistical analysis

#### Univariate analysis

Univariate variable analysis was performed to discover kinematic features that differed (i) between stimulation conditions in cervical dystonia, and between cervical and generalised dystonia. To establish significance, we used either Wilcoxon (when paired between preand postoperatively) or Mann-Whitney U-tests, and report p-values adjusted for multiple comparisons (Benjamini Hochberg false discovery rate correction, FDR). Effect sizes were computed using rank-biserial correlation. To aid interpretation, we ranked variables by their effect sizes. Statistical analyses were done in Python with the statmodels package (0.15.0). Correlation analysis was performed to identify relationships between head angle excursions and annotated scores. Pearson correlations were calculated in Python with the scipy package (1.4.1).

#### Harmonic analysis

The strength of the fundamental tremor frequency and its first harmonic (double the fundamental frequency) were calculated using the distance correlation between their instantaneous phases. The harmonic strengths were determined using the head angles for each axis of motion respectively. Distance correlations were calculated in Python with the dcor package (0.6)

## Results

A total of 88 patients were retrospectively rated in both treatment conditions by three independent raters using the TWSTRS severity rating scale, as well as the TWSTRS-2 tremor item. We ensured that the raters were blinded to the disease and treatment status of the patients. For severity ratings, we focused on the attempted neutral, ‘null’ head position captured in each video, aiming to capture dystonic head deviations in three principal axes: yaw for torticollis, tilt for laterocollis, and pitch for antero-/retrocollis. We observed considerable variation in the annotated scores among the raters, which differed between axes: while clinical raters strongly agreed (mean Cohen-kappa score: 0.86) on torticollis severity, they only moderately agreed on laterocollis and antero-/retrocollis scores (mean Cohen-kappa scores: 0.65, 0.67 respectively) (Figure S2). Across all axes, DBS treatment led to a significant reduction in clinical ratings, i.e., severity (Figure S3A). However, effect sizes differed considerably within each axis: 0.71 for torticollis, 0.49 for laterocollis and 0.85 for anteroretrocollis. Moreover, individual clinical scores exhibited a strong correlation between the pre- and post-operative DBS conditions (Figure S4A). Longitudinally, post-operative clinical ratings in the torticollis and laterocollis directions demonstrated a negative correlation with the duration between pre- and post- operative evaluations, in line with the clinical observation of delayed effects (Figure S4B). However, there was no correlation between the difference in clinical rating from pre- to post-operation and the duration of time (Figure S4C).

We proceeded to assess the clinical relevance of the visual perceptive framework in accurately capturing angular deviations of the head. We extracted the excursion of head angles from attempted neutral head positions for each patient. The head angles strongly agreed with clinical scores for all prinicipal axes of motion (r *≥* 0.66, Figure 2A). We further observed a significant reduction of head angle deviations in each axis by DBS (Figure 2B) with largest effect sizes in torticollis (0.59), followed by laterocollis (0.46) and anteroretrocollis (0.38). To further investigate the relationship between head angle deviations and clinical characteristics, we divided the patients into three phenotypic groups based on their dominant axis of deviation. We discovered that each group of patients exhibited a significant change from pre- to post-operative evaluations only in their respective dominant axis of deviation (effect sizes: torticollis 0.76; laterocollis 0.93; anteroretrocollis 0.60; Figure 2C). For instance, patients with a dominant torticollis excursion only demonstrated a significant change in yaw but not in other axes. Furthermore, we found no systematic excursion in a particular direction for any axis of movement (Figure S5). The pre- and post-operative head angles exhibited a strong correlation (Figure 2D), indicating a reduction in angle excursion following treatment but not a complete elimination. However, we did not observe a systematic correlation between head angles in different axes of motion among both patients and controls (Figure 2E).

**Figure 1:**
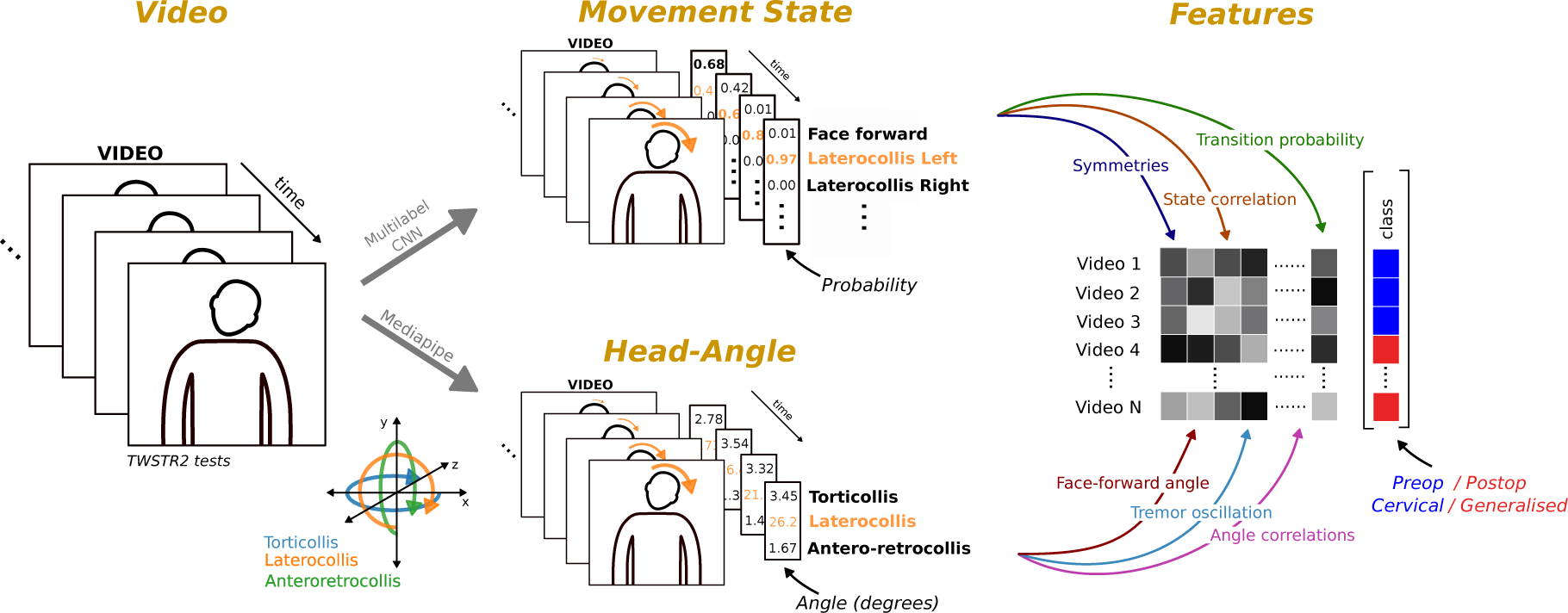
Measurement of static and kinematic features using computer vision workflow. Videos comprising individual frames are fed into convolutional neural network models that predict the movement state, i.e., the probability of the head direction of a patient, and track face-mesh coordinates to derive head angles for each frame. Head angle deviations can be extracted directly during periods of the video where a patient attempts a neutral face forward position. Using the full video, kinematic features can be constructed from the movement states predictions and angles, e.g., the correlation between axis of rotation or dystonic tremors. Features can be stored and compared across groups, such as operation status or disease.

**Figure 2:**
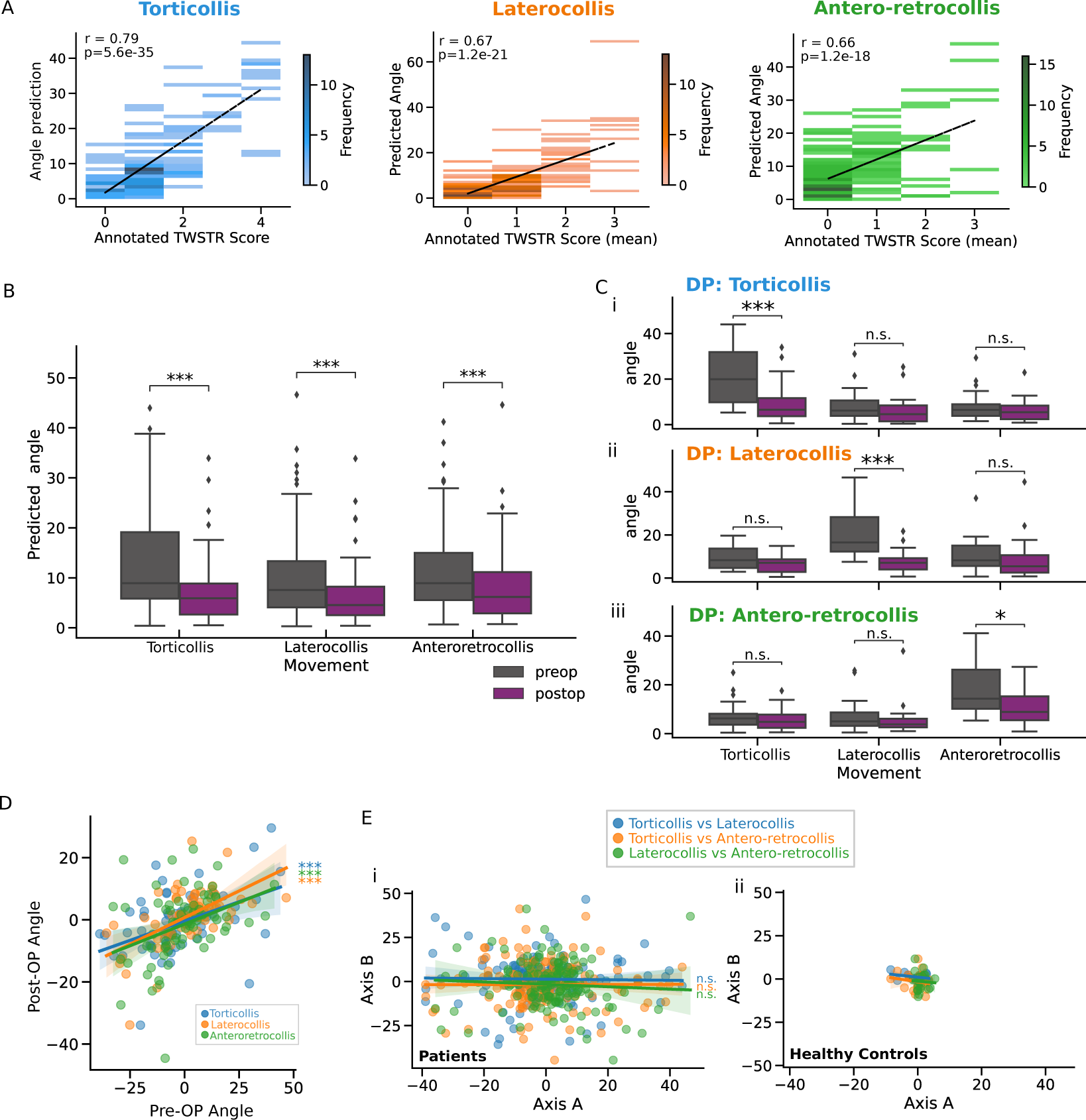
Computer vision analysis of head angle during periods of face-forward. **A** 2-D histograms for comparing video derived head-angle (absolute angle) and clinically assigned TWSTRS scores for each axis of motion. **B** Box plots showing (absolute) pre- (grey) and post- (purple) operative angles, for each axes of movement. Median and interquartile ranges are displayed in each plot. **C** Like (B) but patients are separated into their dominant phenotypes, i.e., their dominant axis of deviation from face-forward. **D** Scatter plot showing correlation of predicted pre- and post-operative head angles for each movement axis. **E** Scatter plots correlating each pair of axes of motion for (i) patients and (ii) healthy controls, for pre- and post-op combined. Correlations were Pearson r tests. Group tests were Mann-Whitney U-tests: * p*<* 0.05; ** p*<* 0.01; *** p*<* 0.001.

Next, we hypothesized that relying solely on the measurement of static head angular deviations is inadequate for providing a comprehensive description of the diverse range of dystonic movement abnormalities observed in real-world clinical assessments. An example of the head-angle kinematics from a full clinical examination both pre- and post-operation is shown in Figure 3A. Therefore, we conducted an explorative analysis of videos encompassing the entire TWSTRS severity assessment, utilising a comprehensive set of clinically inspired kinematic variables (Figure 3C left). First, we find that several kinematic variables are significantly larger pre-operatively. The top five differentiating features included oscillatory characteristics in each axis (ranging from 2-10 Hz) and correlations of movement states. Notably, the effect sizes of these kinematic features were generally larger than those of angle deviations during attempted neutral face-forward positioning, suggesting that they are more responsive to DBS intervention. To identify kinematic features that are predominantly associated with a favourable treatment response, we further divided the sample into responder and non-responder groups based on the degree of improvement in overall clinical rater scores (i.e., patients with *≥* or *<* 30% improvement, Figure 3B). After repeating statistical tests between DBS conditions for the responder and non-responder groups respectively, we found that the top five kinematic features were also more strongly modulated in the responder group (calculated as effect size of responders minus the effect size of non-responders, Figure 3C right). To understand the time-scales at which dystonic movements were modulated by DBS, we applied multiscale entropy analysis to the head-angle timeseries. At DBS ON, patients displayed less complex head movements at shorter timescales (i.e., *<* 1s) (Figure S6), but no significant differences were observed at longer scales (i.e., *>* 1s), indicating that neural circuit interventions restore movement regularity on subsecond time scales.

**Figure 3:**
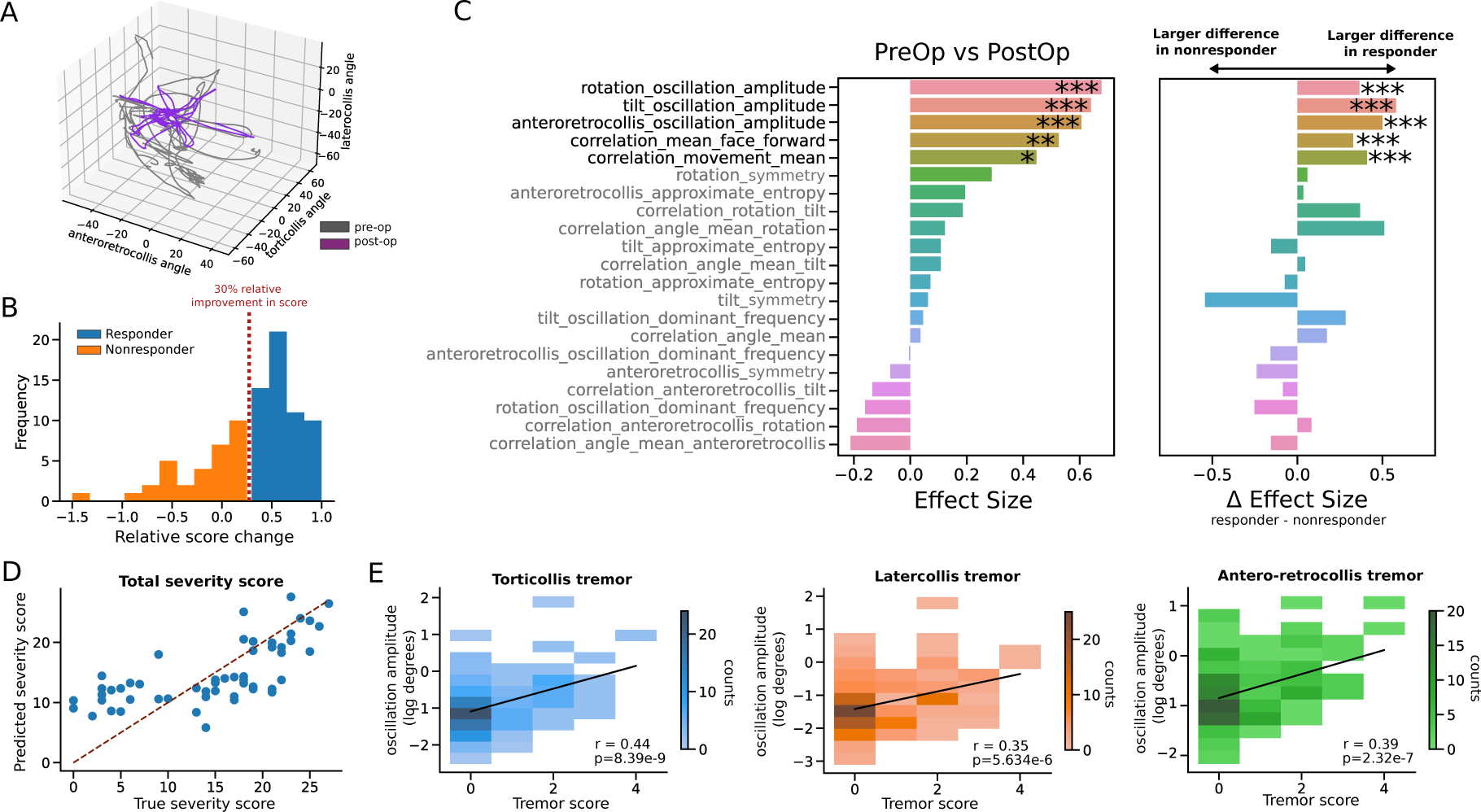
Statistical analysis of kinematic variables from full-videos. Kinematic variables (e.g., head tremor amplitude and frequency, correlations of movement states) were derived from the full-video as patients performed a series of clinically assigned movements. **A** Example of the head-angle kinematics for a randomly chosen patient pre- and post-operation reveals a more structured movement with DBS. **B** Responders are patients who observed a 30% relative improvement in their clinically rated score from pre-operative DBS off to post-operative DBS on. **C** Summary of statistical analysis, showing (i) effect size of Wilcoxon tests between pre- and post-operation (rank-biserial correlation, positive effect indicating variable is larger during pre-operation period) and (ii) the difference in effect sizes of the responder group and non-responder group (all tests Benjamini Hochberg FDR corrected). **D** A scatter plot showing the relationship between predicted values of total severity scores using additive sequential feature selection on a linear model with a combination of kinematic and static features (mean absolute error 4.79). The dotted red line corresponds to line of perfect agreement between predicted and true holistic scores. **E** 2-D histograms for comparing video derived oscillations for each axis of motion and a clinically assigned tremor severity score (not defined by axis of motion). Fitted linear model in black. Significance levels: * p*<* 0.05; ** p*<* 0.01; *** p*<* 0.001.

Despite the original purpose of the scores to capture head-angle deviations from the natural face-forward position, we hypothesized a significant influence of a broader clinical impression beyond pure angular deviations. Hence, we investigated whether the kinematic variables also correlated with the clinically annotated scores. We found that various kinematic features positively and negatively correlated with the scores of each axis of head motion (FDR corrected p-values, Figure S7). These kinematic features included symmetries of movement, correlations of movement states, and oscillation amplitudes and frequencies. By collapsing the TWSTRS sub-item scores into an average, we further defined a holistic, clinical dystonia severity measure. Notably, the correlation strength of kinematic features to the holistic score increased when compared to the scores of each independent axis of motion, in some cases surpassing the correlation strength of the head-angle deviations (Figure S7). To independently verify the holistic score, we collected the original total TWSTRS severity score for a sub-cohort of patients (scores ranging from 0-27). Using a linear model with additive sequential feature selection, we found the optimal model to predict the total TWSTR severity score included a combination of static head-angle deviations and kinematic features (mean absolute error 4.79, Figure 3D). Repeating sequential feature selection with only head-angle deviations produced inferior predictions (mean absolute error 5.63), suggesting that head-angle deviations must be accompanied by kinematic features for the automated assessment of overall dystonic severity. We further examined the oscillatory kinematic features along each axis and found that they correlated with clinically assigned tremor scores (Figure 3E). However, we found no significant correlation between the oscillation amplitudes and face-forward angle deviations (Figure S8), suggesting that they capture distinct dimensions of dystonic movements independent of the angle deviations in static head position.

To assess the robustness and validity of the extended kinematic feature set, we used an independent, out-of-sample data set comprising pre- and post-operative videos of 30 patients with generalised dystonia collected between 2002 and 2008. It should be noted that patients with generalised dystonia tend to also have craniocervical disease manifestations [13]. Due to the video framing, only the upper-half poses of patients were captured, thereby excluding additional signs of generalised dystonia such as twisting in limbs from the analysis (Figure 4A). As clinical ratings and periods of static face-forward were unavailable for the generalised dystonia dataset, only kinematic variables (and not head angle excursions) were extracted. We repeated the statistical analysis of kinematic variables as modulated by DBS in generalised dystonia patients (Figure S9). Remarkably, we observed that the same five dynamic variables that exhibited the strongest response to DBS in cervical dystonia were also significantly modulated in the generalised dystonia patients (Figure S9A).

**Figure 4:**
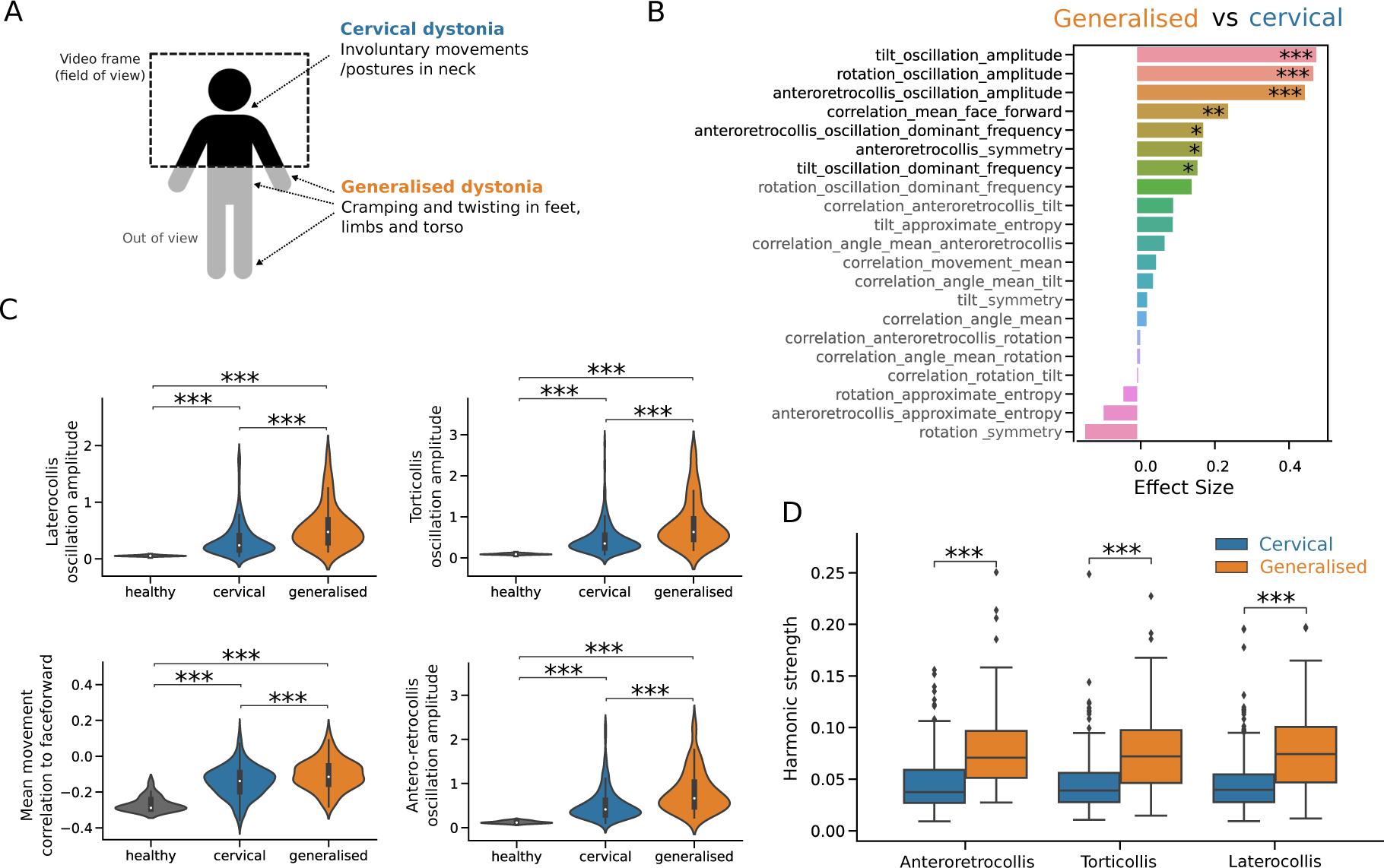
Comparison of generalised and cervical dystonia patients using kinematics variables. Annotations of face-forward periods were unavailable and thus only kinematic variables from the full-videos were extracted. **A** Schematic describing the typical visibility of patient pose captured by videos. Markers indicate common symptoms in cervical and generalised dystonia. **B** Effect sizes (rank-biserial correlation) of Mann-Whitney U-tests (Benjamini-Hochberg FDR corrected) between generalised and cervical dystonia patients (positive effective indicating variable is larger in generalised dystonia patients). **C** Violin plots of variables that are significantly larger in generalised dystonia patients relative to cervical dystonia patients (none were found as statistically significant vice-versa). Healthy controls shown for reference. **D** Comparison of oscillation harmonic strengths between cervical and generalised dystonia patients along each axis of motion. Harmonic strength is measured per patient as the distance correlation between the phases of the dominant tremor frequency and its harmonic (twice the dominant frequency). Mann-Whitney U-tests: * p*<* 0.05; ** p*<* 0.01; *** p*<* 0.001.

Finally, we asked whether kinematic features could differentiate between cervical and generalised dystonia. Analysis within the kinematic feature space revealed seven features that displayed a clear differentiation between cervical and generalised patients (Figure 4B). These features were consistently larger in generalised dystonia patients. Among the significant features, four corresponded to the previously identified five kinematic features that were preferentially modulated by DBS: oscillatory features in all three axes and a mean movement correlation with the face-forward position. Additionally, two frequency-related variables capturing the frequency of head oscillations in the laterocollis and anteroretrocollis axes were also significantly larger in generalised dystonia patients. Notably, all the identified features exhibited pronounced differences compared to healthy controls (Figure 4C), indicating their sensitivity to dystonic movements. Considering the prominent involvement of oscillatory kinematic features, which are associated with tremor, we further examined the strength of harmonics in cervical and generalised dystonia patients. Our analysis revealed that generalised dystonia patients exhibited stronger harmonics in head tremor oscillations across all axes of motion, in comparison to cervical dystonia patients (Figure 4D). Moreover, multiscale entropy analysis showed that generalised dystonia patients displayed maximal entropy at much earlier timescales relative to cervical dystonia patients (Figure S10). This suggests that longer-scale movement patterns hold valuable information to distinguish between different stages of dystonia.

## Discussion

In this study, we developed a visual perceptive framework using convolutional neural networks to comprehensively evaluate dystonia based on clinical video recordings. Our unique dataset comprised longitudinal video documentation of cervical and generalised dystonia patients’ full clinical assessments at multiple medical centers, including those with and without DBS. This enabled us to comprehensively evaluate head movements in both task-constrained, static conditions and quasi-naturalistic, dynamic conditions, providing a holistic assessment of dystonia. Beyond technical validation, we demonstrate the framework’s utility to augment clinical judgement and facilitate insights pertinent to disease states and the readout of neural circuit intervention effects.

Clinical scales, commonly used to assess dystonia and other neurological disorders, have inherent limitations due to clinimetric issues, likely stemming from the oversimplification of complex disease phenomenology into low-dimensional ordinal parameters [9, 11–13]. While necessary in time-sensitive clinical settings, this oversimplification comes at the expense of precision, granularity, and ultimately, ecological validity. An illustration of this is evident when comparing the relatively simple contemporary scoring approaches with Oppenheim’s detailed phenotypical account of dystonia from 1911 [39]

For example, TWSTRS omits some key clinical features of dystonia which only become evident with dynamic, voluntary movements and undoubtedly influence overall clinical judgement. Whilst adaptions to TWSTRS have incorporated tremor related features [40, 41], there is still a growing demand for more objective and granular disease metrics [3, 18]. Already widely used for quantitative phenotyping in experimental neuroscience, computer vision approaches have recently emerged as a promising new tool for clinical assessments [20, 23, 24, 42].

We first demonstrate the robustness and clinical applicability of our deep learning framework in accurately inferring head-angle deviations during attempted ‘null’ head positions from diverse clinical videos captured using consumer-grade hardware. Our visual perceptive approach surpasses various vision-based frameworks that relied on multiple or specialized depth cameras to automate ratings [21, 22], and achieves comparable performance to a recent study by Zhang et al. [20]. However, a distinguishing feature of our approach is the ability to estimate head angles in real-time using a portable device such as smartphones or tablets. This capability enables its practical implementation in clinical point-of-care settings and at-home monitoring, enhancing accessibility and convenience. Moreover, we have applied our framework to diverse clinical videos from multiple centres with slightly differing protocols, showing the effects of neuromodulation in both focal and generalised dystonia, highlighting the generalisability of our tool and specificity of our findings.

The key advantage of our framework lies in its capability to analyse full video examinations of patients. To showcase this, we reverse-engineered complex clinical observations such as dystonic overflow, tremor and the action-dependent dynamics of dystonic movements into objectively measurable kinematic features. Albeit highly informative and clinically indispensable, these dimensions are not explicitly part of the TWSTRS. To this end, we first tuned a convolutional neural network to parse naturalistically occurring 3D head positions into discrete, geometrically defined states. By projecting each sample into a high-dimensional space comprising clinically inspired and interpretable kinematic features, we successfully identified a set of five kinematic variables that exhibited maximal differentiation across therapy states. These were in addition to expected improvements in head-angle deviations, which have been shown in prior studies [7]. In other words, these features demonstrated the most pronounced response to neuromodulation, rendering them highly specific to the behavioral downstream effects of the neural circuit intervention. Furthermore, our analysis revealed that these same features were closely associated with a favorable treatment response to DBS, as defined clinically by a relative score reduction of more than 30% [32, 33, 35]. This finding not only underscores the relevance of these features but also highlights their potential as reliable indicators of effect and efficacy of neural circuit interventions.

Our analysis revealed that three of the kinematic features associated with DBS effect were related to head oscillations. Additionally, multi-scale entropy analysis highlighted that neuromodulation exerts the most profound effects on movement regularity on a subsecond time scale, pointing rather to high frequency phasic than low frequency, tonic aspects of dystonic movements. This finding aligns with recent evidence indicating that head tremor is a prevalent manifestation in the majority of patients with cervical dystonia [14, 43, 44]. The recognition of tremor as a core clinical characteristic only recently led to the inclusion of a quantifiable tremor item in the revised version of the TWSTRS [9]. Tremor-related features emerged most consistently across contrasts, strongly highlighting the previously less well documented role of oscillatory aspects in dystonia pathophysiology and therapy. Notably, tremor has been associated with impaired physical functioning and pain, which are crucial dimensions of quality of life in dystonia [45, 46]. Therefore, the linkage between kinematic features and patient-centered outcomes provides an avenue for further investigations into ‘disease architectures’ comprised by multiple phenotyping axes. The remaining correlational features we identified in our analysis provide further insights into potential manifestations of dystonic overflow and multiaxial involvement, as expressed in an abnormal covariance of head movement trajectories. These features were evaluated throughout dynamic movement trajectories, capturing a key characteristic of dystonia, namely the provocation of involuntary, dystonic movements through voluntary action. In an independent validation dataset comprising 30 generalised dystonia patients, we found the same kinematic features reflected pallidal DBS effects, confirming aforementioned results in cervical dystonia patients. This demonstrates the generalisability of our findings to different states of disease progression and further reinforces their disease-specific nature. Furthermore, experimental investigations in rodent models of dystonia suggest that similar correlational features are independent predictors of genetic susceptibility factors in rodent models of dystonia, establishing a first hint for their neurobiological and translational relevance [15].

To gain further insights into the discriminatory potential of these kinematic features, we attempted to distinguish different disease states, namely focal-cervical and generalised dystonia, within the kinematic feature space. A total of seven features exhibited significant differences, with four of the previously identified kinematic features among them. Notably, control patients exhibited the lowest values, followed by cervical and then generalised dystonia patients. Multi-scale entropy analysis further highlighted that focal and generalised dystonias show a pronounced difference of regularity in both subsecond and longer time scales *>* 1 second, pointing to a more profound dysfunction of motor control in generalised dystonia. Overall, these observations align well with the concept of a dystonic phenotypical continuum wherein severity progressively increases [13], and suggests that these kinematic features sensitively capture disease state and progression, which is of critical relevance for interventional studies. Moreover, the emergence of a unified feature set specific to both cervical and generalised dystonia aligns with recent findings demonstrating the convergence of a multisynaptic neural network underlying both dystonia subgroups [47]. The observed motor behavioural disorganisation is mirrored on the neurobiological level by pathologically irregular neuronal firing patterns associated with the dystonic state [15, 48], overall suggesting that kinematic features can be a powerful readout of brain circuit function.

To better understand what information the kinematic features captured, we next correlated them with clinically annotated scores and measured angular head deviations. Despite the clinically annotated scores purposed to capture natural head-angle deviations from attempted null position, we found various dynamic features that were correlated with clinical, but not head-angle deviations. This included clinical scores along each axis but also a holistic severity score. These results imply that the kinematic features are, at least partially, encoded by different neurobiological substrates. In terms of oscillatory features, this finding aligns with recent work on diverging symptom-specific circuit components for tremor versus dystonia [49]. Moreover, it suggests that clinicians may unintentionally incorporate more complex kinematic aspects from a patient’s dystonic symptomatology into their clinical scores to more accurately reflect the global clinical impression. This could explain some of the discussed limitations of current scoring approaches, which may be confounding factors for score-based therapeutic or brain-behaviour association studies. Within the context of rapidly emerging adaptive neurotechnologies [50] and connectomic neuroimaging techniques [47], intriguing use cases for our deep learning approach come to mind, such as pathophysiologically motivated circuit interrogation or guidance of adaptive and personalized neuromodulatory treatment regimes[50, 51].

Our study has several limitations that should be considered. Firstly, our assessments were limited to videos focusing on the upper body, thereby neglecting dystonic phenomena occurring in other regions. However, it is important to note that cervical dystonia is one of the most common forms of dystonia, and studying head kinematics provides a valuable entry point for investigating digital pathosignatures of dystonia, given the relative simplicity of head movements compared to whole-body movements. Secondly, although our measurements of oscillation amplitudes demonstrated substantial clinical validity, it is important to note that the degree of validity was slightly lower compared to previous investigations that exclusively focused on oscillations occurring in the head’s null position [52]. We deliberately opted to derive tremor amplitudes from the full videos, considering that tremor in cervical dystonia exhibits variation in relation to head position [43, 53]. This approach allowed for a more ecologically valid estimation of tremor but also introduced natural variability into the measurements. Thirdly, we did not incorporate information on DBS parametrization. The location of the implanted lead and the electrical stimulation fields are known to be important predictors of therapy response in dystonia [35, 47, 54]. This omission may have influenced the performance of our kinematic features in capturing therapy state contrasts, as suboptimal responses could reduce the overall distance between therapy states in the feature space. To partially address this limitation, we conducted a subgroup analysis specifically focusing on clinically determined good responders.

Overall, these findings highlight the potential of our visual perceptive framework to enhance and augment dystonia diagnosis, monitoring and therapy by uncovering consistent latent pathosignatures. The proposed modern vision-based approach expands upon traditional principles of ‘medical cinematography’ in movement science. Video-derived kinematic pathosignatures may not only inform neural circuit therapeutics but also address the critical need for objective and standardized evaluation methods in the form of digital biomarkers. Their high sensitivity has recently been shown to facilitate clinical trials, genotype predictions and continuous monitoring in neurological disorders [24, 42, 55]. Moreover, our framework may bridge methodological gaps between clinical and experimental neuroscience, which has already widely adapted computer vision for phenotyping animal models of dystonia [15–17]. We envisage the proposed tool to strengthen translational and precision medicine approaches in modern neurology.

## Data Availability

All data produced in the present study are available upon reasonable request to the authors.

## Acknowledgements

RP, SRS, CWI, JV, AK, AS acknowledge the Deutsche Forschungsgemeinschaft (DFG, German Research Foundation) Project-ID 424778381-TRR 295 (A01, A06, B04, C01). MF and CWI acknowledge the Interdisciplinary Center for Clinical Research, IZKF, IZKF-Z2-CSP13, A-421, S-506 at the University Hospital Wuerzburg. SRS is a Fellow of the Thiemann Foundation. The project was partially funded by the Arbeitskreis Botulinumtoxin e.V. (to CWI and MF), Merz Pharma and Ipsen Pharma (to CWI). CWI is funded by the VERUM foundation. JV receives funding from the European Union’s Horizon 2020 research and innovation programme under the EJP RD COFUND-EJP N° 825575 (EurDyscover).

## Author contributions

Conceptualisation: CWI; Methodology: RLP, MF, CWI; Software: RLP, MM; Resources: CWI, JV, CS, JK, AS, MW, AKH, SP, AK, IMS, WE, JM, CM, MR Analysis: RLP, MF, CWI, DZ, LF, MM, SRS; Writing - Original Draft: RLP, MF; Writing - Review: RLP, MF, SRS, CWI; & Editing: RLP, MF, CWI, MM, SRS, MR;

## Supplementary Material

### S1 Model variable interpretation

Below we provide more details on the interpretation of the derived variables from the deep learning framework:

#### Head angles

- Angle torticollis: Head-angle deviation from face-forward in yaw axis when a patient is sitting in a neutral position. Positive angle = right, Negative angle = left.
- Angle laterocollis: Head-angle deviation from face-forward in tilt axis when a patient is sitting in a neutral position. Positive angle = right tilt, Negative angle = left tilt.
- Angle antero/retrocollis: Head-angle deviation from face-forward in antero/retrocollis axis when a patient is sitting in a neutral position. Positive angle = anterocollis, Negative angle = retrocollis.

#### Correlation features

- Correlation movement mean: Mean correlation coefficient between all predicted movement states.
- Correlation mean face forward: Mean correlation coefficient of each movement state to faceforward movement state.

#### Head oscillations

- Oscillation amplitude: The amplitude of the largest peak in a Fourier transform of the angles. For each axis respectively.
- Oscillation frequency: The frequency of the largest peak in a Fourier transform of the angles. For each axis respectively.

#### Symmetry features

- Symmetry rotation: Proportion of time head was oriented in one direction compared to the opposite direction for the rotation states (left or right).
- Symmetry tilt: Proportion of time head was oriented in one direction compared to the opposite direction for the tilt states (left or right).
- Symmetry anteroretrocollis: Proportion of time head was oriented in one direction compared to the opposite direction for the antero/retrocollis states (forward or backward).

### S2 Supplementary figures

**Figure S1:**
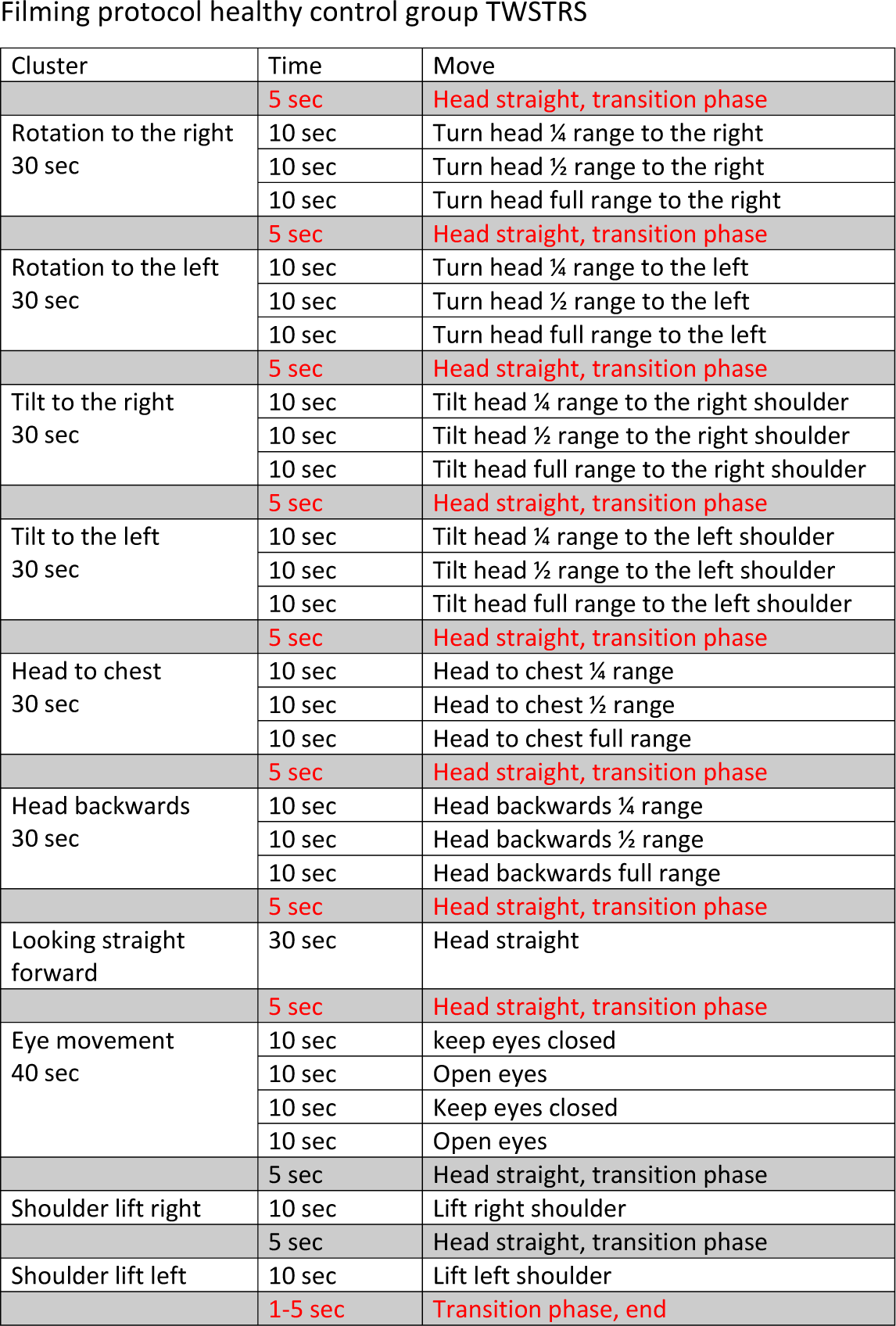
Healthy controls filming protocol.

**Figure S2:**
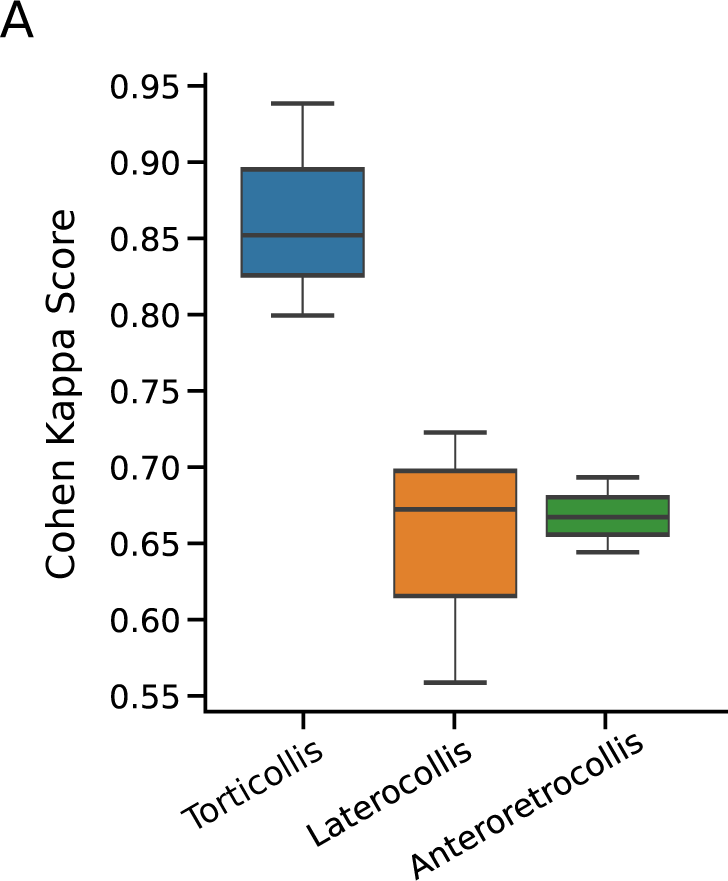
Agreement of clinical raters. Distribution of Cohen-Kappa-scores by axis (3-paired comparisons per axis).

**Figure S3:**
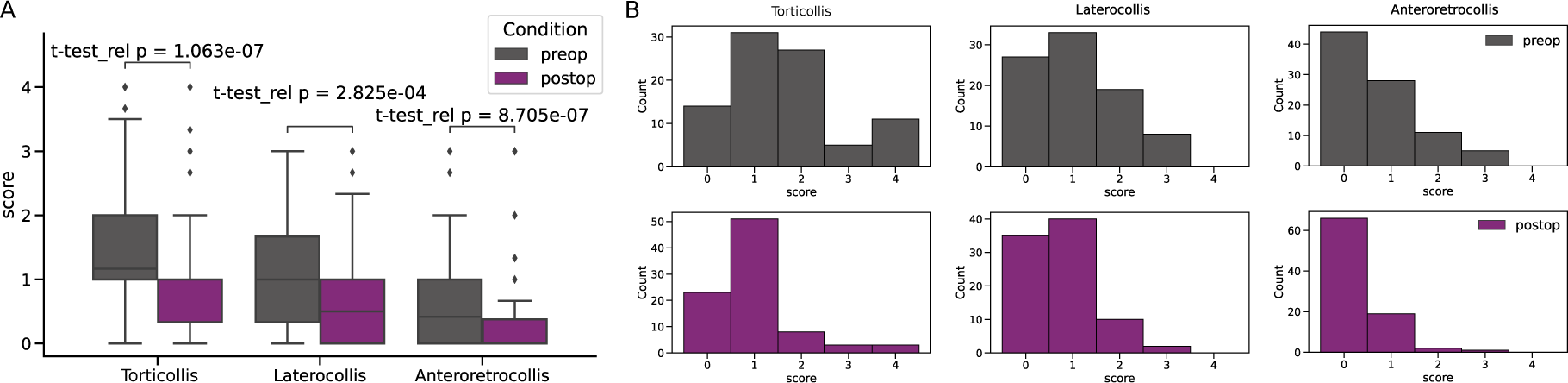
Distribution of pre- and post-operative clinically annotated scores. **A** Box plots showing clinically rated patient scores (mean across clinical raters) for pre- (grey) and post- (purple) operation, for each axes of movement. Median and interquartile ranges are displayed in each plot. **B** Pre- (top) and post- (bottom) operative distributions of scores. Maximum score is 4 for torticollis and 3 for laterocollis and anteroretrocollis.

**Figure S4:**
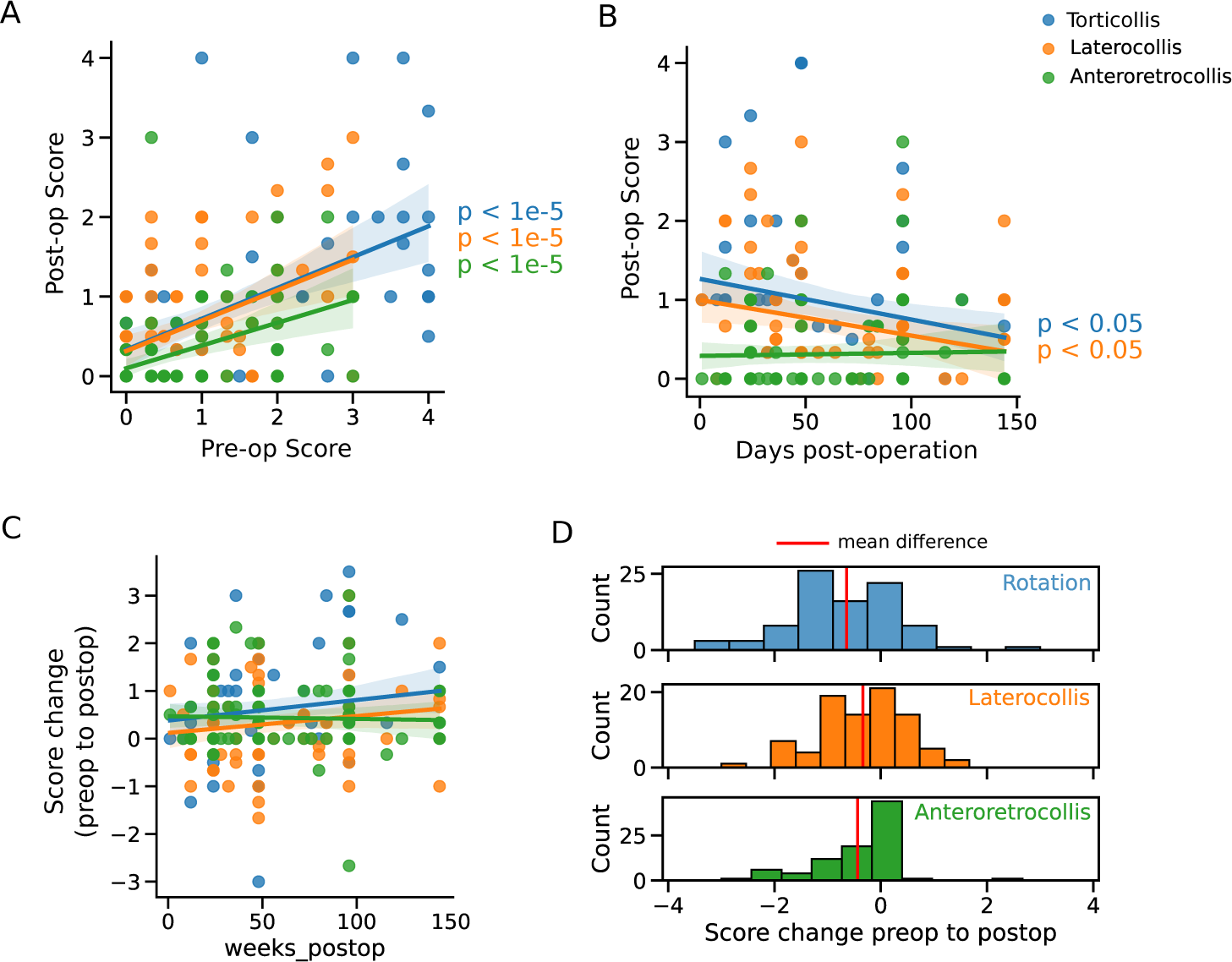
Comparison of pre- and post-operation scores. **A** Scatter plot showing correlations of pre- and post-operation scores. Fitted linear model. **B** Correlation between date of post-operative assessment and score. **C** A scatter plot of the relative change in clinicallly assigned scores between pre- and post-op (pre-op score subtracted from post-op score). **D** Histograms of relative changes in scores (pre-op score subtracted from post-op score). By colours: torticollis (blue), laterocollis (orange) and anteroretrocollis (green)

**Figure S5:**
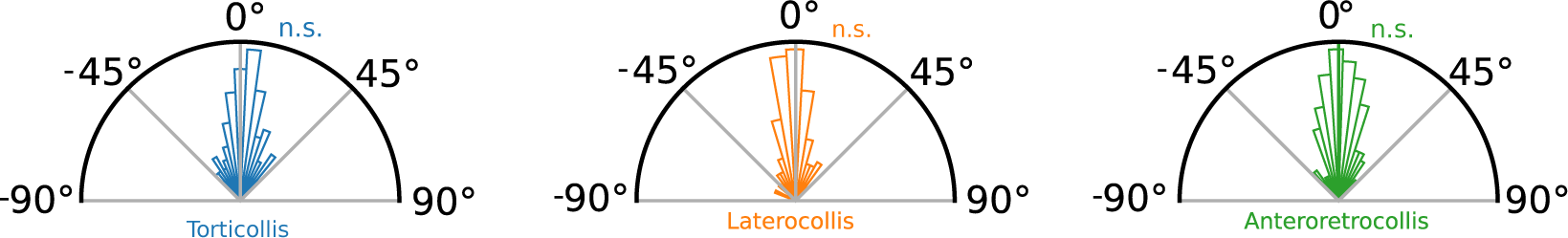
Deviations from face-forward are centred at zero at the group-level. Polar histograms showing that the average angle does not significantly deviate in any direction. One-sample t-tests were non-significant in each axis of motion.

**Figure S6:**
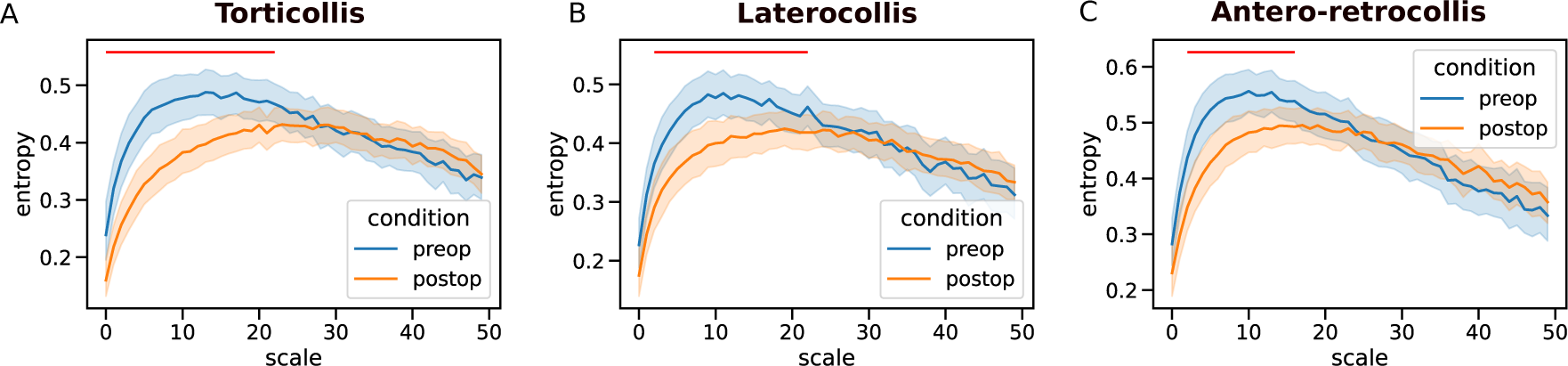
Multiscale entropy reveals only short timescale differences pre- to post-operation. Multiscale entropy analysis (approximate entropy) is applied to the each head-angle time-series for increasing scales (Python EntropyHub 0.2). Scale (x-axis) is defined in units of video frames (videos were standardised to a sampling rate of 25 frames per second). Maximal entropy is observed earlier for pre-operative recordings relative to post-operation. Red lines indicate scales with significant differences between pre and post-operation (paired t-test, p*<* 0.05).

**Figure S7:**
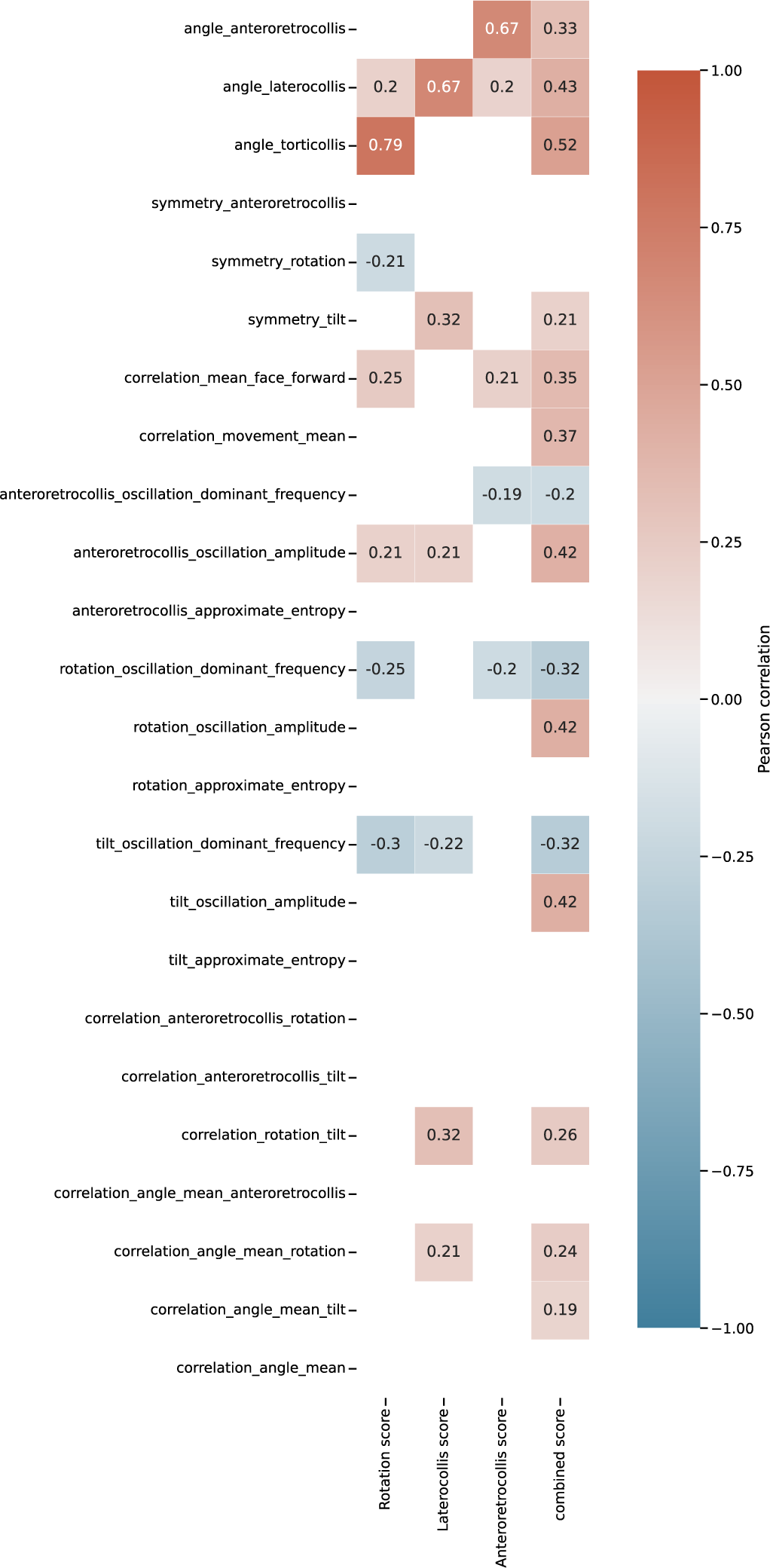
Correlation of kinematic features with annotated scores. Clinically annotated scores (mean across raters) for head-angle deviations from neutral face-forward along each axis are correlated with engineered kinematic features from full videos. A holistic score taken as the mean clinical rating across the three axes is also correlated with kinematic features. Only significant (FDR corrected, *p <* 0.05) correlations are shown.

**Figure S8:**
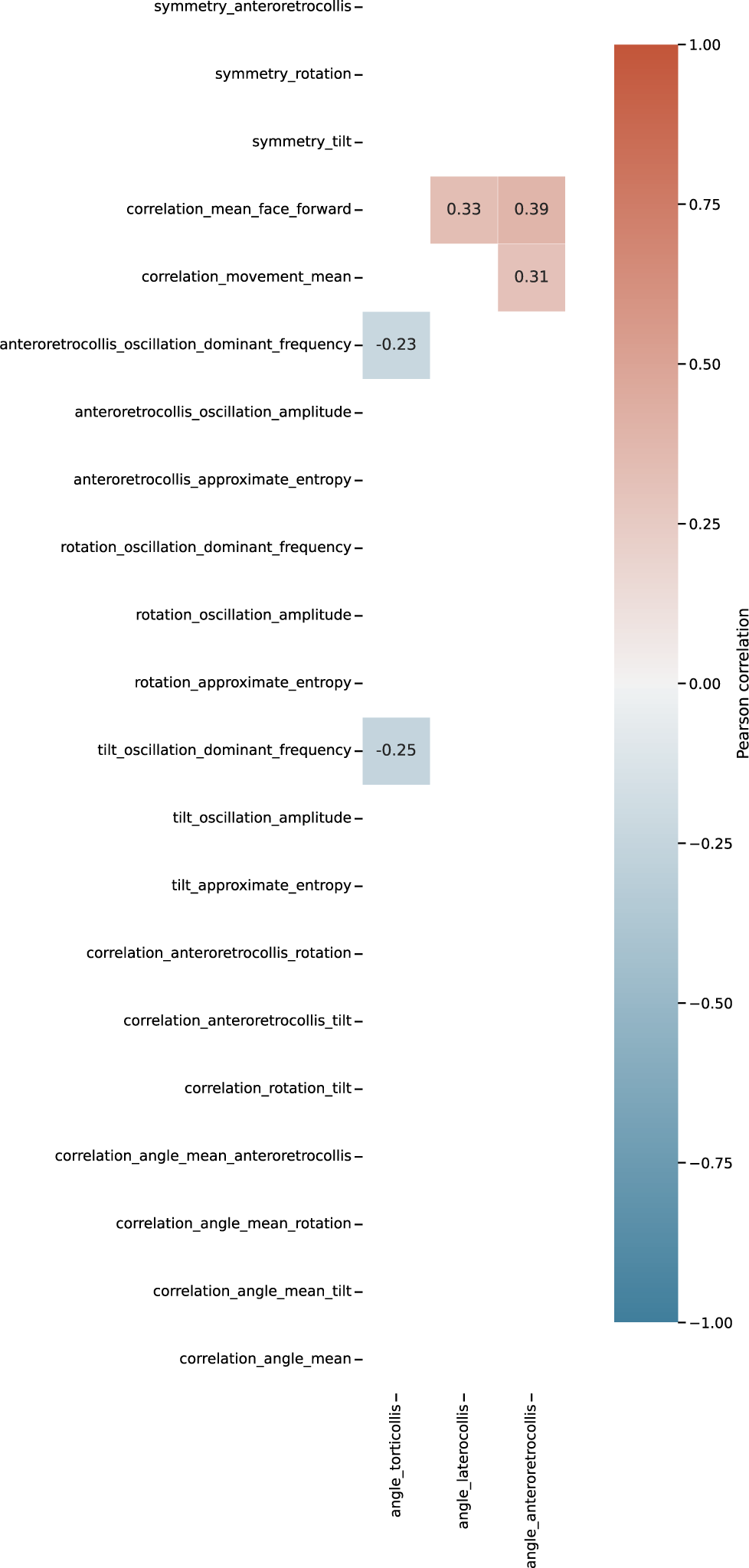
Correlation of kinematic features with face-forward angles. Head-angle deviations during attempted neutral face-forward are correlated with engineered kinematic features from full videos. Only significant (FDR multiple comparisons corrected, *p <* 0.05) correlations are shown.

**Figure S9:**
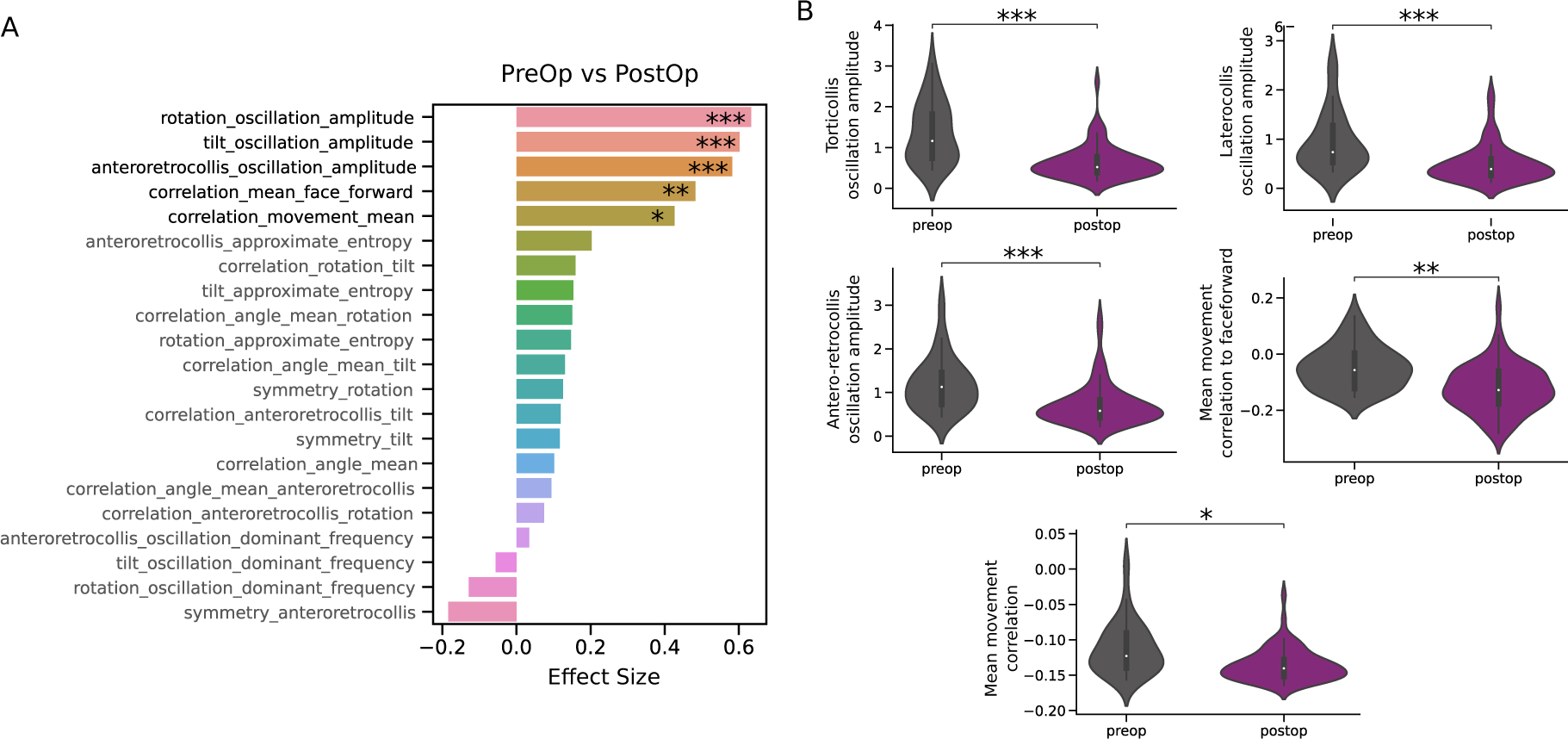
Validation of full video kinematics in cohort of generalised dystonia patients. **A** Effect size (rank-biserial correlation) of dynamical variables between pre- and post-operation with Wilcoxon tests. **B** Violin plots of variables that are significantly larger pre- (grey) relative to post- (purple) operation. Significance levels: * p*<* 0.05; ** p*<* 0.01; *** p*<* 0.001.

**Figure S10:**
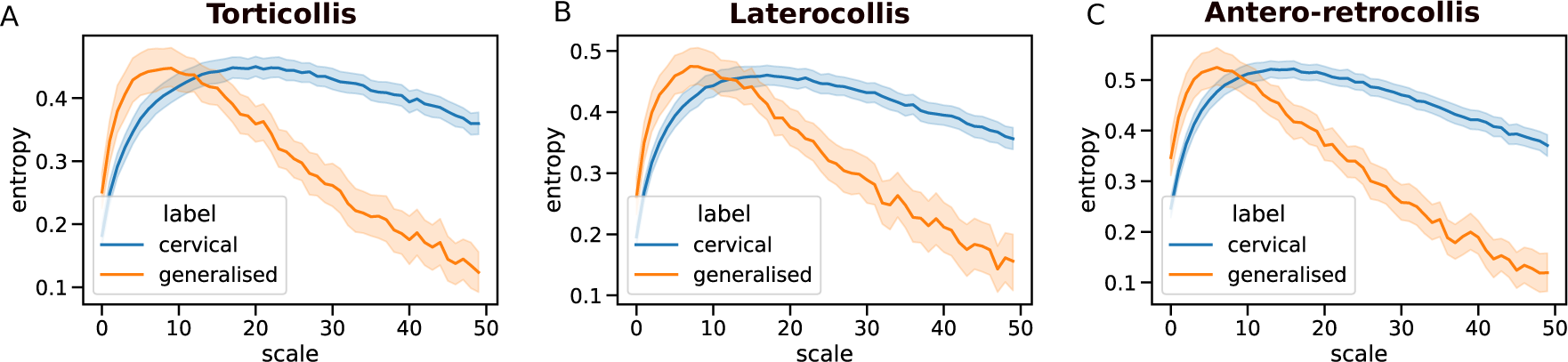
Multiscale entropy reveals differences between generalised and cervical patients at all scales. Multiscale entropy analysis (approximate entropy) is applied to the each head-angle time-series for increasing scales (Python EntropyHub 0.2). Maximal entropy is observed at earlier scales for generalised dystonia patients.

## References

1. Albanese, A. et al. Phenomenology and classification of dystonia: a consensus update. eng. Movement Disorders: Official Journal of the Movement Disorder Society 28, 863–873. issn: 1531-8257 (June 2013).

2. Balint, B. et al. Dystonia. en. Nature Reviews Disease Primers 4. Number: 1 Publisher: Nature Publishing Group, 1–23. issn: 2056-676X. https://www.nature.com/articles/s41572-018-0023-6 (Sept. 2018).

3. Kilic-Berkmen, G. et al. The Dystonia Coalition: A Multicenter Network for Clinical and Translational Studies. Frontiers in Neurology 12, 660909. issn: 1664-2295. https://www.ncbi.nlm.nih.gov/pmc/articles/PMC8060489/ (Apr. 2021).

4. De, A., et al. Machine Learning in Tremor Analysis: Critique and Directions. Movement Disorders 38, 717–731. https://movementdisorders.onlinelibrary.wiley.com/doi/abs/10.1002/mds.29376 (2023).

5. Schreglmann, S. R. et al. Non-invasive suppression of essential tremor via phase-locked disruption of its temporal coherence. Nature communications 12, 363 (2021).

6. Lalli, S. & Albanese, A. The diagnostic challenge of primary dystonia: evidence from misdiagnosis. eng. Movement Disorders: Official Journal of the Movement Disorder Society 25, 1619– 1626. issn: 1531-8257 (Aug. 2010).

7. Blahak, C. et al. Improvement of head and neck range of motion induced by chronic pallidal deep brain stimulation for cervical dystonia. Journal of Neural Transmission 128, 1205–1213 (2021).

8. Comella, C. L. et al. Teaching tape for the motor section of the Toronto Western Spasmodic Torticollis Scale. eng. Movement Disorders: Official Journal of the Movement Disorder Society 12, 570–575. issn: 0885-3185 (July 1997).

9. Comella, C. L. et al. Clinimetric Testing of the Comprehensive Cervical Dystonia Rating Scale. Movement disorders : official journal of the Movement Disorder Society 31, 563–569. issn: 0885-3185. https://www.ncbi.nlm.nih.gov/pmc/articles/PMC4833533/ (Apr. 2016).

10. Burke, R. E. et al. Validity and reliability of a rating scale for the primary torsion dystonias. eng. Neurology 35, 73–77. issn: 0028-3878 (Jan. 1985).

11. Comella, C. L., et al. Rating scales for dystonia: a multicenter assessment. eng. Movement Disorders: Official Journal of the Movement Disorder Society 18, 303–312.issn: 0885-3185 (Mar. 2003).

12. Krystkowiak, P. et al. Reliability of the Burke-Fahn-Marsden scale in a multicenter trial for dystonia. eng. Movement Disorders: Official Journal of the Movement Disorder Society 22, 685–689. issn: 0885-3185 (Apr. 2007).

13. Albanese, A. et al. Dystonia rating scales: critique and recommendations. Movement disorders : official journal of the Movement Disorder Society 28, 874–883. issn: 0885-3185. https://www.ncbi.nlm.nih.gov/pmc/articles/PMC4207366/ (June 2013).

14. Shaikh, A. G. et al. Dystonia and Tremor: A Cross-Sectional Study of the Dystonia Coalition Cohort. eng. Neurology 96, e563–e574. issn: 1526-632X (Jan. 2021).

15. Knorr, S. et al. The evolution of dystonia-like movements in TOR1A rats after transient nerve injury is accompanied by dopaminergic dysregulation and abnormal oscillatory activity of a central motor network. eng. Neurobiology of Disease 154, 105337. issn: 1095-953X (July 2021).

16. Rauschenberger, L. et al. Peripheral nerve injury elicits microstructural and neurochemical changes in the striatum and substantia nigra of a DYT-TOR1A mouse model with dystonia-like movements. eng. Neurobiology of Disease 179, 106056. issn: 1095-953X (Apr. 2023).

17. Brown, A. M. et al. Cerebellar Dysfunction as a Source of Dystonic Phenotypes in Mice. eng. *Cerebellum (London*, England). issn: 1473-4230 (July 2022).

18. Contarino, M. F. et al. Unmet Needs in the Management of Cervical Dystonia. Frontiers in Neurology 7. issn: 1664-2295. https://www.frontiersin.org/articles/10.3389/fneur.2016.00165 (2016).

19. Vanmechelen, I. et al. Assessment of movement disorders using wearable sensors during upper limb tasks: A scoping review. Frontiers in Robotics and AI 9, 1068413 (2023).

20. Zhang, Z. et al. Hold that pose: capturing cervical dystonia’s head deviation severity from video. eng. Annals of Clinical and Translational Neurology 9, 684–694. issn: 2328-9503 (May 2022).

21. Ye, C. et al. Pilot Feasibility Study of a Multi-View Vision Based Scoring Method for Cervical Dystonia. Sensors (Basel, Switzerland) 22, 4642. issn: 1424-8220. https://www.ncbi.nlm.nih.gov/pmc/articles/PMC9230118/ (June 2022).

22. Nakamura, T. et al. Pilot feasibility study of a semi-automated three-dimensional scoring system for cervical dystonia. eng. PloS One 14, e0219758. issn: 1932-6203 (2019).

23. Friedrich, M. U. et al. Smartphone video nystagmography using convolutional neural networks: ConVNG. en. Journal of Neurology. issn: 1432-1459. 10.1007/s00415-022-11493-1 (Nov. 2022).

24. Morinan, G. et al. Computer vision quantification of whole-body Parkinsonian bradykinesia using a large multi-site population. npj Parkinson’s Disease 9, 10 (2023).

25. Tien, R. N. et al. Deep learning based markerless motion tracking as a clinical tool for movement disorders: Utility, feasibility and early experience. Frontiers in Signal Processing 2. issn: 2673-8198. https://www.frontiersin.org/articles/10.3389/frsip.2022.884384 (2022).

26. Esteva, A. et al. Deep learning-enabled medical computer vision. en. npj Digital Medicine 4. Number: 1 Publisher: Nature Publishing Group, 1–9. issn: 2398-6352. https://www.nature.com/articles/s41746-020-00376-2 (Jan. 2021).

27. Seethapathi, N. et al. Movement science needs different pose tracking algorithms arXiv:1907.10226 [cs, q-bio]. July 2019. http://arxiv.org/abs/1907.10226.

28. Colyer, S. L. et al. A Review of the Evolution of Vision-Based Motion Analysis and the Integration of Advanced Computer Vision Methods Towards Developing a Markerless System. Sports Medicine - Open 4, 24. issn: 2198-9761. 10.1186/s40798-018-0139-y (June 2018).

29. Hammadi, Y. et al. Evaluation of Various State of the Art Head Pose Estimation Algorithms for Clinical Scenarios. en. Sensors 22. Number: 18 Publisher: Multidisciplinary Digital Publishing Institute, 6850. issn: 1424-8220. https://www.mdpi.com/1424-8220/22/18/6850 (Jan. 2022).

30. Lugaresi, C. et al. MediaPipe: A Framework for Building Perception Pipelines arXiv:1906.08172 [cs]. June 2019. http://arxiv.org/abs/1906.08172.

31. Baltrusaitis, T., Robinson, P. & Morency, L.-P. *OpenFace: An open source facial behavior analysis toolkit* en. in 2016 *IEEE Winter Conference on Applications of Computer Vision (WACV)* (IEEE, Lake Placid, NY, USA, Mar. 2016), 1–10. isbn: 978-1-5090-0641-0. http://ieeexplore.ieee.org/document/7477553/.

32. Volkmann, J. et al. Pallidal deep brain stimulation in patients with primary generalised or segmental dystonia: 5-year follow-up of a randomised trial. eng. The Lancet. Neurology 11, 1029–1038. issn: 1474-4465 (Dec. 2012).

33. Volkmann, J. et al. Pallidal neurostimulation in patients with medication-refractory cervical dystonia: a randomised, sham-controlled trial. eng. The Lancet. Neurology 13, 875–884. issn: 1474-4465 (Sept. 2014).

34. Kupsch, A. et al. Pallidal deep-brain stimulation in primary generalized or segmental dystonia. eng. The New England Journal of Medicine 355, 1978–1990. issn: 1533-4406 (Nov. 2006).

35. Reich, M. M. et al. Probabilistic mapping of the antidystonic effect of pallidal neurostimulation: a multicentre imaging study. eng. Brain: A Journal of Neurology 142, 1386–1398. issn: 1460-2156 (May 2019).

36. Meng, F. et al. Procrustes: A python library to find transformations that maximize the similarity between matrices. Computer Physics Communications 276, 108334 (2022).

37. Keshmiri, S. Entropy and the Brain: An Overview. Entropy 22, 917. issn: 1099-4300. https://www.ncbi.nlm.nih.gov/pmc/articles/PMC7597158/ (Aug. 2020).

38. Fagerholm, E. D. et al. A primer on entropy in neuroscience. en. Neuroscience & Biobehavioral Reviews 146, 105070. issn: 0149-7634. https://www.sciencedirect.com/science/article/pii/S0149763423000398 (Mar. 2023).

39. Klein, C. & Fahn, S. Translation of Oppenheim’s 1911 paper on dystonia. eng. Movement Disorders: Official Journal of the Movement Disorder Society 28, 851–862. issn: 1531-8257 (June 2013).

40. Comella, C. et al. Reliability of the Severity subscale of the revised Toronto Spasmodic Torticollis Rating Scale (TWSTRS-2) (S15.001). en. Neurology 84. Publisher: Wolters Kluwer Health, Inc. on behalf of the American Academy of Neurology Section: April 21, 2015. issn: 0028-3878, 1526-632X. https://n.neurology.org/content/84/14_Supplement/S15.001 (Apr. 2015).

41. Comella, C. L. et al. Clinimetric testing of the comprehensive cervical dystonia rating scale. Movement Disorders 31, 563–569 (2016).

42. Alty, J. et al. The TAS Test project: a prospective longitudinal validation of new online motor-cognitive tests to detect preclinical Alzheimer’s disease and estimate 5-year risks of cognitive decline and dementia. BMC Neurology 22, 266. issn: 1471-2377. 10.1186/ s12883-022-02772-5 (July 2022).

43. Shaikh, A. G., Zee, D. S. & Jinnah, H. A. Oscillatory head movements in cervical dystonia: Dystonia, tremor, or both? eng. Movement Disorders: Official Journal of the Movement Disorder Society 30, 834–842. issn: 1531-8257 (May 2015).

44. Hvizdosová, L. et al. The Prevalence of Dystonic Tremor and Tremor Associated with Dystonia in Patients with Cervical Dystonia. en. Scientific Reports 10. Number: 1 Publisher: Nature Publishing Group, 1436. issn: 2045-2322. https://www.nature.com/articles/s41598-020-58363-2 (Jan. 2020).

45. Vu, J. P. et al. Head tremor and pain in cervical dystonia. eng. Journal of Neurology 268, 1945–1950. issn: 1432-1459 (May 2021).

46. Junker, J. et al. Quality of life in isolated dystonia: non-motor manifestations matter. Journal of neurology, neurosurgery, and psychiatry, jnnp–2020–325193. issn: 0022-3050. https://www.ncbi.nlm.nih.gov/pmc/articles/PMC8356023/ (Feb. 2021).

47. Horn, A. et al. Optimal deep brain stimulation sites and networks for cervical vs. generalized dystonia. eng. Proceedings of the National Academy of Sciences of the United States of America 119, e2114985119. issn: 1091-6490 (Apr. 2022).

48. Darbin, O. et al. An Entropy-Based Model for Basal Ganglia Dysfunctions in Movement Disorders. BioMed Research International 2013, 742671. issn: 2314-6133. https://www.ncbi.nlm.nih.gov/pmc/articles/PMC3671275/ (2013).

49. Paoli, D. et al. DBS in tremor with dystonia: VIM, GPi or both? A review of the literature and considerations from a single-center experience. Journal of Neurology 270, 2217–2229. issn: 0340-5354. https://www.ncbi.nlm.nih.gov/pmc/articles/PMC10025201/ (2023).

50. Neumann, W.-J. et al. Adaptive Deep Brain Stimulation: From Experimental Evidence Toward Practical Implementation. Movement disorders (2023).

51. Hollunder, B. et al. Toward personalized medicine in connectomic deep brain stimulation. en. Progress in Neurobiology 210, 102211. issn: 0301-0082. https://www.sciencedirect.com/science/article/pii/S0301008221002252 (Mar. 2022).

52. Vu, J. P. et al. Head tremor in cervical dystonia: Quantifying severity with computer vision. eng. Journal of the Neurological Sciences 434, 120154. issn: 1878-5883 (Mar. 2022).

53. Albanese, A. & Sorbo, F. D. Dystonia and Tremor: The Clinical Syndromes with Isolated Tremor. eng. *Tremor and Other Hyperkinetic Movements (New York*, N.Y.) 6, 319. issn: 2160-8288 (2016).

54. Lange, F. et al. Machine versus physician-based programming of deep brain stimulation in isolated dystonia: A feasibility study. Brain Stimulation 16, 1105–1111 (2023).

55. Kadirvelu, B. et al. A wearable motion capture suit and machine learning predict disease progression in Friedreich’s ataxia. en. Nature Medicine 29. Number: 1 Publisher: Nature Publishing Group, 86–94. issn: 1546-170X. https://www.nature.com/articles/s41591-022-02159-6 (Jan. 2023).

